# Ultrasensitive C-reactive protein as biomarker of cardiometabolic risk in a group of obese children and adolescents

**DOI:** 10.1101/2020.08.14.20175182

**Authors:** Giovanna Lúcia Oliveira Bonina Costa, Bianca Carolina da Silva Medeiros, Yara Nobre Araújo, Leandro Silva Menezes Júnior, Paulo Uendel da Silva Jesus, Carlos Alberto Menezes

## Abstract

**Objective:** To establish the importance of evaluating ultrasensitive C-reactive protein (us-CRP) in a pediatric group with obesity as the main biomarker, detecting, as early as possible, cardiometabolic complications.

**Methods:** This is a control-case, cross-sectional study involving the biochemical and anthropometric evaluation of 342 children and adolescents participating in the Preventive Medicine Service, in Aracaju, Sergipe, Brazil. When evaluated anthropometrically, it was observed that, in 235 of the cases, the body mass index (BMI) above the 97th percentile or the Z-score greater than +2 allowed their classification as obese. The control group consisted of 107 non-obese individuals. The sample was divided into three age groups according to the International Diabetes Federation (FID): 6–10 years, 10–16 years and >16 years, representing 45%, 39% and 14% of the sample population, respectively.

**Results:** The CRP-us showed an average value of 2.36 ± 1.28 mg/dL in the obese group, while in the control group, the result found was 0.01 ± 0.1 mg/dL. There was a significant correlation between the increase in CRP levels in the obese group and other biochemical and anthropometric findings in the same group, such as: reduced HDL, elevated triglycerides, higher BMI, and increased abdominal circumference (AC). Homocysteine, in turn, proved to be a biomarker with little specificity in the sample in question.

**Conclusion:** The ultra-sensitive C-reactive protein, already fundamentally correlated with increased cardiovascular risk in adults, demonstrates to be a validated biomarker, showing high sensitivity even in pediatric obese populations.

**RESUMO:** *Objetivo:* Estabelecer a importância da avaliação da proteína C reativa ultrassensível (PCR-us) como principal biomarcador em um grupo pediátrico com obesidade, detectando precocemente possíveis complicações cardiometabólicas.

*Métodos:* Trata-se de um estudo caso-controle, transversal envolvendo avaliação bioquímica e antropométrica de 342 crianças e adolescentes, participantes do Serviço de Medicina Preventiva, em Aracaju-Sergipe, Brasil. Quando avaliados antropometricamente, observou-se que em 235 participantes o IMC acima do percentil 97 ou o Z-escore maior que +2 permitia classificá-los como obesos. O grupo controle consistiu de 107 indivíduos não obesos. A amostra foi dividida em três faixas etárias de acordo com a Federação Internacional de Diabetes (FID): 6–10 anos, 10–16 anos e > 16 anos, representando, respectivamente, 45%, 39% e 14% da população amostral.

*Resultados:* A PCR-us apresentou um valor médio de 2.36 ± 1.28 mg/dL no grupo obeso, enquanto no grupo controle o resultado encontrado foi de 0.01 ± 0.1 mg/dL. Observou-se uma correlação significativa do aumento dos níveis de PCR-us no grupo obeso com outros achados bioquímicos e antropométricos do mesmo conjunto, como: redução do HDL, elevação de triglicérides, maior grau de IMC e aumento da CA. A homocisteína, por sua vez, demonstrou ser um biomarcador com pouca especificidade na amostra em questão.

*Conclusão:* A proteína C reativa ultrassensível, já fundamentadamente correlacionada ao aumento do risco cardiovascular em adultos, demonstra ser um validado biomarcador, apresentando alta sensibilidade mesmo em populações pediátricas obesas.

## INTRODUCTION

Since the 1970s, pediatric overweight and obesity remain broadly in growth throughout the world. In this phase, children can be strongly affected, both by adult behavior and by genetic factors, which contribute to the variation in fat accumulation and, therefore, body composition^1,2^. This sharp growth may be responsible for the increased cardiometabolic risk during childhood.

From 1975 to 2016, world trends about the change in Body Mass Index (BMI) show a global increase, per decade, of 0.32kg/m² in girls and 0.40kg/m² in boys. This results in an overall increase from 0.7% to 5.6% and from 0.9% to 7.8% in the female and male genders, respectively, in this time interval^3^. However, there is a noticeable stabilization in pediatric obesity growth in developed countries and its maintenance in currently developing countries^4,5^. In Brazil, the Family Budget Research (FBR), in partnership with the Ministry of Health (MH), showed that 33.5% of children of ages between 5–9 years old were overweight, whereas 16.6% of the boys and 11.8% of the girls were obese; considering the age range of 10–19 years, the overweight was of 21.7% in the male gender and 19.4% in the female gender, the Southeast being the region with the highest overweight levels, and the South, the one with the highest increase in its frequency^6,7^.

Obesity is considered a complex and multifactorial chronic disease, the result of a long period of positive energy balance, in which are involved genetic and environmental factors, as well as an associated low-level inflammatory process^8,9^. Currently, there is evidence that the adipose tissue is not only energy storage but also acts as an active endocrine organ, which operates in many ways in organic systems, contributing to the inflammatory process in obese individuals. By raising cytokine secretion, the abnormal values of metabolites as lipids, fatty acids, and cytokines released by the adipose tissue activate monocytes and increase inflammatory response, which stimulates macrophages and adipocytes to produce proinflammatory adipokines, such as leptin tumor necrosis factor-alpha (TNF-alpha), Interleukin (IL-1), Interleukin-6 (IL-6) and procoagulant substances. Among these, IL-6 stimulates the liver to produce C-reactive protein (CRP), and regulates the production of adiponectin by the adipose tissue^10^. This hormone, antagonistically to the other adipokines, raises the insulin sensibility in tissues and acts as an anti-inflammatory protein hormone. The rise in CRP, synthesized in the liver in many clinical conditions, as infection, trauma, and inflammation, results in significant direct and indirect systemic alterations. In this process, there is a downregulation in nitric oxide (NO), a rise in low-density lipoprotein (LDL-c), facilitating the action of proatherosclerotic genes, adiponectin inhibition, vasoconstriction and stimulus to the platelet adhesion and activation, which may result in occlusion and thrombus formation. Thus, besides being an inflammatory marker of atherosclerosis and coronary events, the CRP is also a disease mediator due to its contribution to lesion formation^8,10,11^.

Once the role of inflammation in obesity development and atherosclerotic disease was determined, different inflammatory biomarkers were searched and identified to add prognostic value to cardiovascular risk, even in the pediatric group. However, world literature is not unanimous regarding the correlation between ultrasensitive CRP (CRP-us) and the risk of infantile-juvenile obesity^12^.

Due to heterogeneity in CRP-us researches as a cardiometabolic risk biomarker, this study’s objective was to establish the importance in CRP-us evaluation in specific obese pediatric groups as the central biomarker, which aims to detect possible early cardiometabolic complications in their pediatric age range.

## MATERIAL AND METHODS

### Sample

This is a transversal, case-control study, with a sample of 342 individuals between 6 and 18 years old, 235 being obese children and adolescents (128 males and 107 females), and 107 of the control group being non-obese children and adolescents (52 males and 55 females). Due to sample heterogeneity, the individuals were divided into three age ranges according to the International Diabetes Federation (IDF): 6–10, 10–16, and over 16 years old.

The study was developed in the Preventive Medicine Service (SEMPRE) in Aracaju –Sergipe, Brazil, constituted by a multi-professional team composed of an endocrinologist, a psychologist, a social assistant, a physical educator, and a nutritionist. The purpose was to follow obese children and adolescents, as well as to offer assistance to parents and family. This work was approved by the Research Ethics Committee in Santa Cruz State University (UESC – Bahia/Brazil), via Brazil Platform, over the consent number 04065412.600005526.

### Anthropometric and biochemical evaluations

Initially, the following anthropometric parameters were assessed on the individuals: weight, height, BMI, abdominal circumference (AC), and arterial pressure (AP). For weight verification, an anthropometric scale was used (Filizola, Brazil). The characterization of obesity was based on the World Health Organization’s BMI charts, following the comparison of age and gender^30^. Thus, the individuals were considered obese when BMI value was above the 97th percentile or the Z-score calculus over +2. The abdominal circumference was measured on the midpoint between the inferior costal border and the iliac crest superior border, and arterial pressure was measured using cuffs with circumference and width appropriate to the age range.

Subsequently, the groups were biochemically evaluated through fasting blood glucose concentration (FBG), insulin, Homeostasis model assessment of insulin resistance (HOMAIR), aspartate aminotransferase (AST), alanine aminotransferase (ALT), gammaglutamyltransferase (GGT), uric acid, total cholesterol (TC), Low-density lipoprotein (LDL-c), High-density lipoprotein (HDL), triglycerides (TG), complete blood count, thyroid-stimulant hormone (TSH), free thyroxine (free T4), insulin-like growth factor (IGF-1), urinary cortisol, homocysteine biomarkers, and CRP-us.

### Laboratory cardiometabolic risk biomarkers measurement

The CRP-us was dosed by the immunoturbidimetry method using the Immage system (Beckman Coulter, USA). The values utilized to conduct the cardiovascular risk stratification (CRS) were: low CRS: < 0.1 mg/dL, medium CRS: 0.1 – 0.3 mg/dL, and high CRS: > 0.3 mg/dL.The total plasmatic homocysteine was determined by the chemiluminescence method, with reference value below 15 mcmol/L. The plasmatic insulin was also determined by the chemiluminescence method, the reference value being below 28.4 mU/mL.

The HOMA-IR (Homeostasis model assessment of insulin resistance) was calculated by multiplying the fasting insulin (mU/mL) by the fasting blood glucose (mg/dL), all divided by 405. The used cutoff value for the characterization of insulin resistance was equal or above 3.16.

### Statistical Analysis

The results were statistically analyzed by the Mann Whitney test, adopting a p< 0.05 as a significance level. The results were obtained through the average, standard deviation, and confidence level. For the statistical analysis, the Graph Pad Prisma software 5.1 was used.

## RESULTS

342 children and adolescents were evaluated, 235 being from the obese group (128 males and 55 females), and 107 individuals being from the non-obese group (52 males and 55 females). The mid-interval of chronologic ages was of 10+2.3 years old for both sexes.

Anthropometric and biochemical data from the male obese group compared to the control group according to the age range are shown in Table 1. It was observed that, in all evaluated age ranges, the obese individuals showed a significant change of anthropometric parameters, with elevated BMI and arterial pressure compared to the control group (6–16 years: P<0.0001; > 16 years: p<0.001). Furthermore, it was verified that these individuals had dyslipidemia with a significant rise of TC (P<0.001) in all the age ranges, a rise of LDL-c, TG, and a decrease of HDL-c, the most prominent differences being found in the 16-year-olds’ range (p<0.001). Regarding the glycemic profile, the individuals over 16 years-old showed fasting blood glucose above the control group (p<0.05). The results also showed that CRP-us was increased in all evaluated groups (p<0.0001).

**TABLE 1.** Anthropometric and biochemical data in a group of male children and adolescents according to age range, control group (n = 52), and obesity (n = 128).

|  |  | 6 to 10 years | 10 to 16 years | > 16 years |
| --- | --- | --- | --- | --- |
| **BMI (Kg/m<sup>2</sup>)** | Control | 17.74 ± 0.99 | 19.92 ± 0.60 | 18.75±0.19 |
|  | Obese | 26.21 ± 0.49*** | 29.11 ± 0.59*** | 33.90±1.37** |
| **AC (cm)** | Control | 64.39 ± 1.28 | 67.30 ± 0.72 | 70.5 ± 0.5 |
|  | Obese | 84.00 ± 1.17*** | 96.07 ± 1.83*** | 105.8 ± 3.92** |
| **FBG (mg/dL)** | Control | 85.78 ± 1.57 | 84.90 ± 1.18 | 88.25 ± 2.75 |
|  | Obese | 86.92 ± 1.23 | 87.61 ± 1.25 | 99.90 ± 1.18* |
| **TC (mg/dL)** | Control | 157.0 ± 5.13 | 156.7 ± 5.56 | 126.8 ± 16.2 |
|  | Obese | 180.1 ± 4.29** | 175.4 ± 3.45** | 211.3 ± 8.59** |
| **LDL (mg/dL)** | Control | 92.72 ± 4.55 | 102.4 ± 7.10 | 93.75 ± 6.25 |
|  | Obese | 109.1 ± 4.31*** | 110.8 ± 4.04*** | 138.8 ± 14.3** |
| **HDL (mg/dL)** | Control | 49.61 ± 1.81 | 45.30 ± 1.85 | 47.50 ± 2.5 |
|  | Obese | 39.02 ± 1.22*** | 37.98 ± 1.15*** | 37.10 ± 0.86** |
| **TG (mg/dL)** | Control | 77.11 ± 6.72 | 90.40 ± 10.5 | 71.00 ± 6.0 |
|  | Obese | 147.0 ± 1.96*** | 136.7 ± 7.7*** | 235.5 ± 27.5** |
| **CRP-us (mg/dL)** | Control | 0.06 ± 0.01 | 0.06 ± 0.01 | 0.01 ± 0.01 |
|  | Obese | 1.43 ± 0.12*** | 2.33 ± 0.17*** | 2.82 ± 0.35*** |

BMI (Body Mass Index), AC (Abdominal Circumference), FBG (Fasting Blood Glucose), TC (Total Cholesterol), LDL-c (Low-density lipoprotein), HDL (High-density lipoprotein), CRP-us (C-Reactive Protein), TG (triglycerides). \*p<0.05, \*\*p<0.001 and \*\*\*p<0.0001 compared to the control group.

Anthropometric and biochemical variations from the obese female groups compared to the control group according to the age range are shown in Table 2. It is noticeable that the anthropometric parameters BMI and AC were significantly elevated in the obese children group compared to the control group, with higher differences in age ranges up to 16 years-old (p<0.0001). Regarding the lipid profile, the obese group TC was increased in the individuals between 6 and 10 years-old (p<0.0001) and 10 to 16 years-old (p<0.001), and increased LDL-c only in the obese group from 6 to 10 years-old (p<0.05). HDL was found to be reduced in the obese groups of 6 to 10 (p<0.0001) and 10 to 16 (p<0.05) years-old, the TG was increased in the obese children under 16 years-old (p<0.0001). Similarly, to the male gender, increased levels of FBS were only observed in obese individuals over 16 years-old (p<0.05). CRP-us was increased in all the age ranges comparing the obese to the control group, with major differences under the 16 years-old range (p<0.0001).

**TABLE 2:** Anthropometric and biochemical data in a group of female children and adolescents according to age range, control group (n = 55), and obesity (n = 107).

|  |  | 6 to 10 years | 10 to 16 years | > 16 years |
| --- | --- | --- | --- | --- |
| **BMI (Kg/m<sup>2</sup>)** | Control | 17.99 ± 0.84 | 17.91 ± 0.41 | 22.5 ± 0.1 |
|  | Obese | 25.39 ± 0.45*** | 29.11 ± 0.71*** | 30.40 ± 1.13* |
| **AC (cm)** | Control | 62.27 ± 2.01 | 65.75 ± 1.01 | 70.0 ± 0.1 |
|  | Obese | 82.97 ± 1.29*** | 90.02 ± 1.33*** | 96.50 ± 3.17* |
| **FBG (mg/dL)** | Control | 83.93 ± 2.07 | 85.67 ± 1.54 | 88.5 ± 0.1 |
|  | Obese | 84.74 ± 1.35 | 87.38 ± 1.03 | 96.00 ± 2.04* |
| **TC (mg/dL)** | Control | 128.1 ± 4.37 | 141.5 ± 3.80 | 120.2 ± 0.1 |
|  | Obese | 172.7 ± 4.35*** | 168.8 ± 4.45** | 182.3 ± 4.23 |
| **LDL (mg/dL)** | Control | 99.73 ± 7.11 | 96.00 ± 3.52 | 99.0 ± 0.1 |
|  | Obese | 110.7 ± 4.04* | 104.1 ± 3.68 | 107.3 ± 5.4 |
| **HDL (mg/dL)** | Control | 58.87 ± 2.33 | 45.76 ± 1.41 | 48.0 ± 0.1 |
|  | Obese | 39.0 ± 0.96*** | 38.2 ± 1.20* | 39.75 ± 0.5 |
| **TG (mg/dL)** | Control | 74.93 ± 4.04 | 81.95 ± 3.21 | 110 ± 2.0 |
|  | Obese | 152.4 ± 8.67*** | 158.9 ± 8.8*** | 159.0 ± 7.4 |
| **CRP-us (mg/dL)** | Control | 0.039 ± 0.009 | 0.04 ± 0.009 | 0.02 ± 0.1 |
|  | Obese | 1.85 ± 0.15*** | 2.27 ± 0.37*** | 2.92 ± 0.14* |

BMI (Body Mass Index), AC (Abdominal Circumference), FBG (Fasting Blood Glucose), TC (Total Cholesterol), LDL-c (Low-density lipoprotein), HDL (High-density lipoprotein), CRP-us (C-Reactive Protein), TG (triglycerides). \* $p < 0.05$ , \*\* $p < 0.001$ and \*\*\* $p < 0.0001$ compared to the control group.

The results referring to the laboratory evaluation with all samples of the obese compared to the control group are shown in Table 3. It can be seen that the obesity group showed altered biochemical variables such as uric acid, insulin, HOMA-IR, TC, HDL, LDL-c, TG, AST, ALT, and GGT. However, it wasn’t verified differences in hormonal dosage between obese and control groups, besides the leukogram not showing a significant difference in the evaluated groups.

**TABLE 3:**
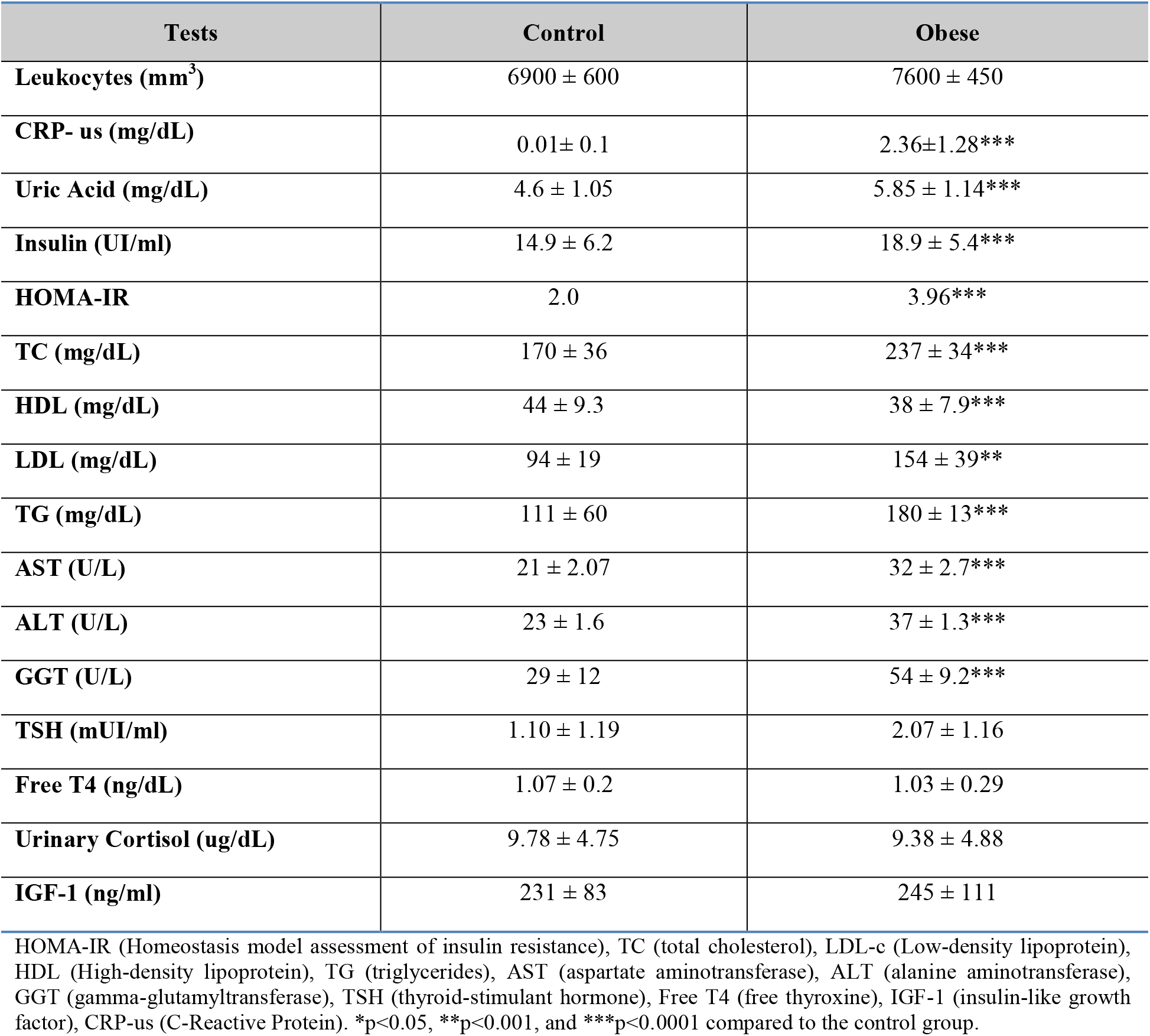
Anthropometric and biochemical data in a group of children and adolescents from both sexes according to age range, control group (n = 107), and obesity (n = 235).

CRP-us and homocysteine levels in the obese and control group, according to sex and age range, are shown in Figure 1. The results revealed that the obese group shows higher CRP-us levels, in all the age ranges and both sexes, when compared to the control group. Such statistical differences were quite significant in males under 16 years-old (p<0.0001). It was not possible to detect a statistical difference in homocysteine levels between obese and control groups.

**Figure 1.**
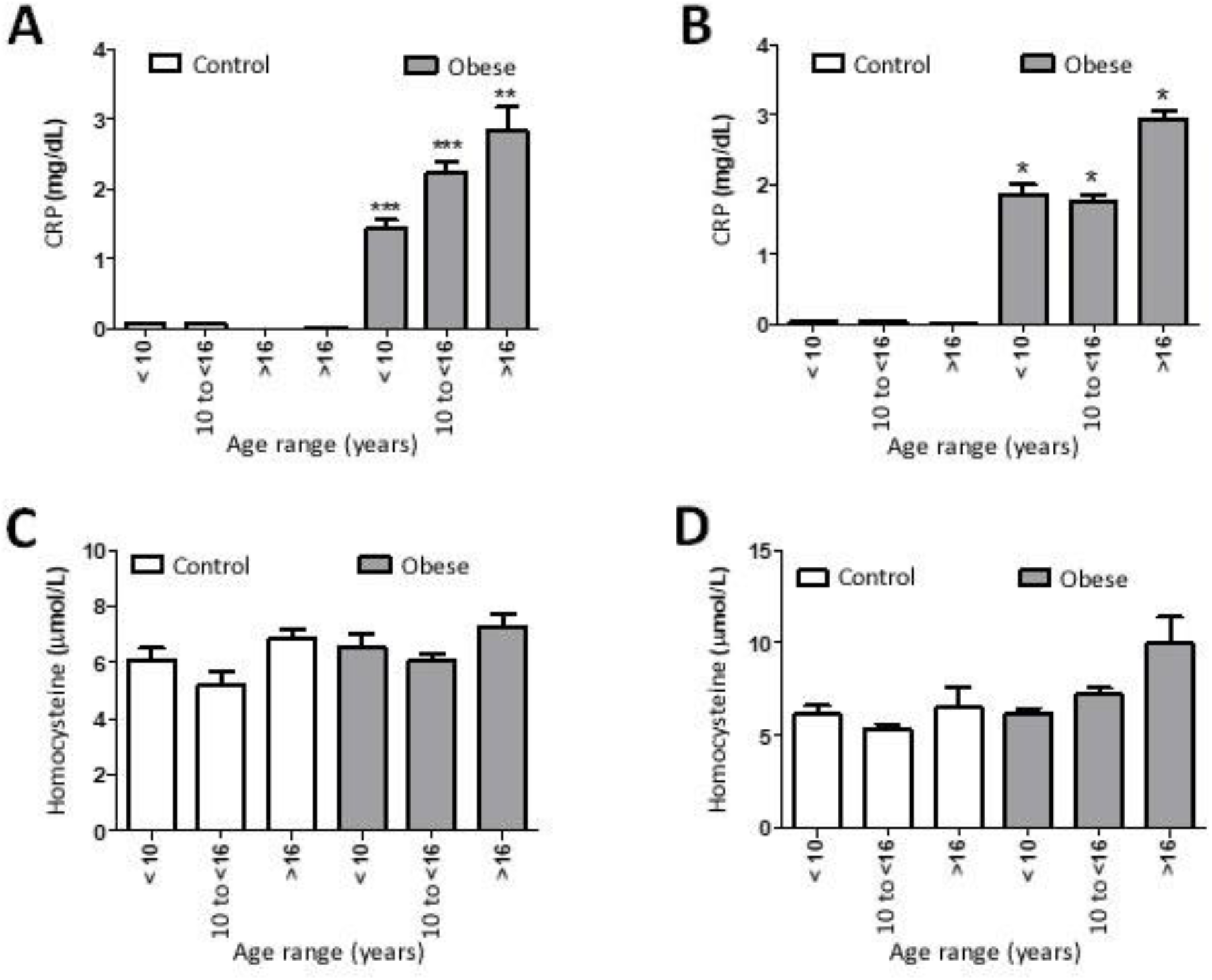
Representation of homocysteine and CRP-us behavior regarding control and obese groups of males (A and C), and females (B and D), according to each studied age range. *p <0.05, **p<0.01, *** p<0.0001.

A comparison between anthropometric variables, BMI and AC and laboratory variables, triglycerides, and HDL, of both sample groups is shown in Figure 2. The representation demonstrates significant differences in CRP-us levels in the obese group with HDL decrease (p = 0.001), triglycerides increase (p = 0.001), higher BMI (p = 0.001), and AC increase (p = 0.001).

**FIGURE 2.**
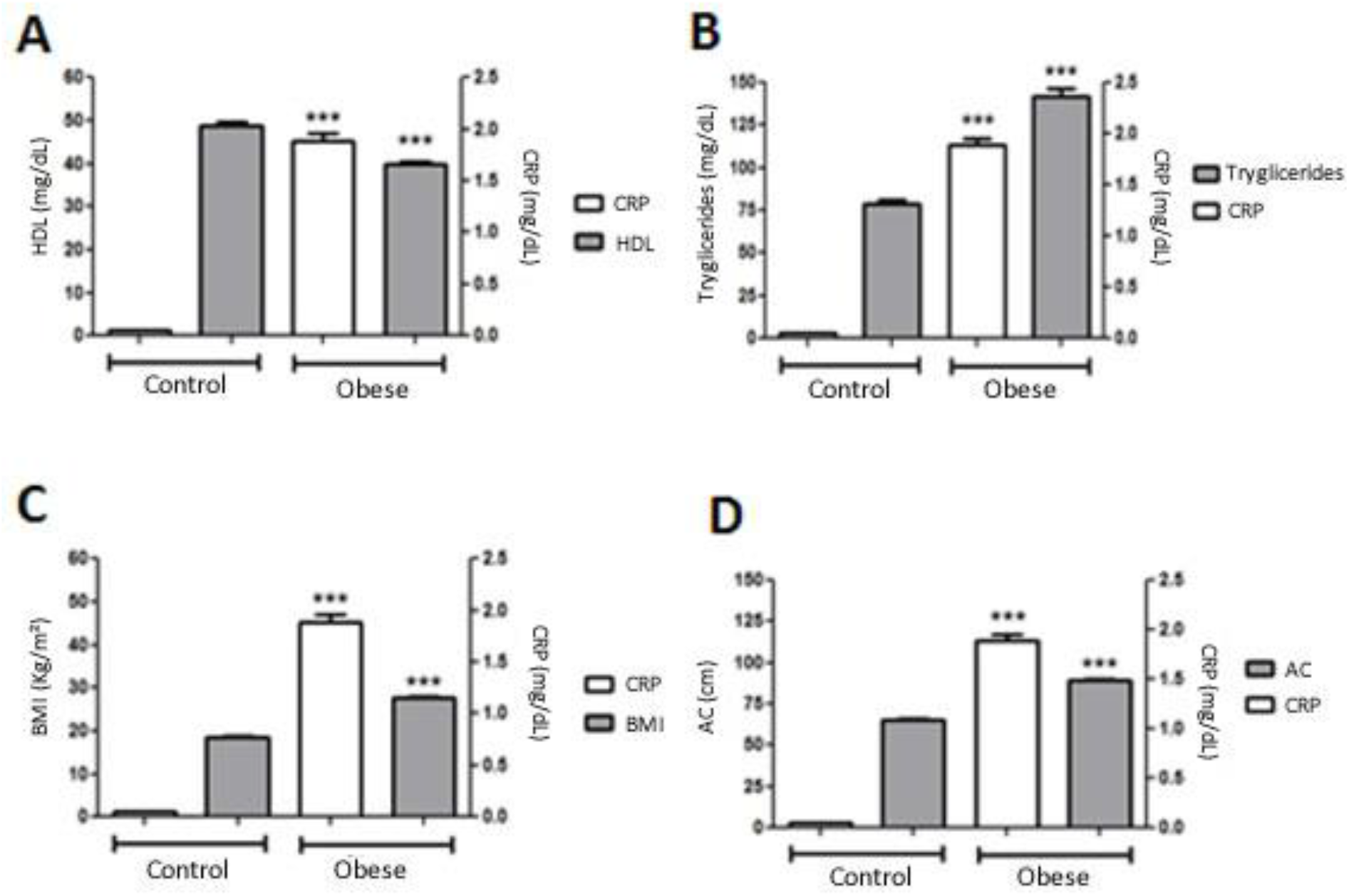
Representation of CRP-us behavior compared to HDL (A), triglycerides (B), BMI (C), and AC (D) variables, in obese and control groups (n = 342, ***p<0.0001).

## DISCUSSION

A biomarker indicates a normal biological process, a pathogenic process, or a pharmacology response to treatment^13^, thus being a quantifiable measure of biologic homeostasis that defines what is normal, and providing subsidies to predict or detect alterations. It is used to identify patients with a high risk of unfavorable outcomes, according to Marshall et al.^12^ When talking about obesity in the pediatric age range, there is no consensus in the world literature regarding the presence of a biomarker and cardiometabolic comorbidities, although, in the adult population, this correlation is already demonstrated^13^. However, studies revealed that high levels of CRP-us are found in obese children and adolescents^14,15,16^. This shows a relationship between obesity and the beginning of an inflammatory process in the obese pediatric population compared to the ones with normal weight^14,15,16^. The results in this study corroborate these findings; however, this increase was more expressive in both sexes in obese individuals over 16 years old compared to non-obese control groups, as shown in Figure 1. This fact is probably due to the higher level of obesity and, consequently, to higher BMI, once that literature relates to the increase of CRP-us to being directly proportional to the BMI level^17^. In a study conducted by Konstantinos Kitsios et al.^18^, the levels of CRP-us were significantly higher in overweight and obese groups compared to control groups. Suhet et al.^19^, when evaluating 350 children, reported a significant association between CRP-us and overweight, believing that the increase of CRP-us may be a potential mediator between obesity and the beginning of atherosclerosis during childhood. A relevant fact in our study was the utilization of the case-control method, pairing both sexes with chronologic age, according to the International Diabetes Federation. This showed with greater clarity the importance of this biomarker in obesity, even in a pediatric group.

The CRP-us is an inflammatory biomarker validated as a cardiovascular risk predictor in apparently healthy adult individuals. The Chronic Disease Control Center (CDC) and the American Heart Association (AHA) classify, according to the relative cardiometabolic risk category, the following CRP-us levels in low cardiac risk (<0.1 mg/dL), medium cardiac risk (0.1–0.3 mg/dL), and high cardiac risk (>0.3 mg/dL). These values were considered to the American and European populations^20^. Using these criteria in this study, it can be seen that obese children and adolescents had higher CRP-us levels in all age ranges. This result allowed the adoption of dietary and behavioral measures to produce a life change for obese individuals. Therefore, it was necessary to apply the effort of a multi-professional team composed of an endocrinologist, a physical educator, a nutritionist, and a psychologist, all of whom stimulated the weight loss and change of habits in these children and their families^21^. This proposition was also adopted by Isasi et al.^22^

Concerning the homocysteine, this cardiometabolic biomarker did not show to have much correlation with obesity in the assessed sample. This fact corroborates what was described by Anderson et al.^23^ that observed the increase of homocysteine in obese patients, although in adults. Another study showed the association between homocysteine and obesity, but concluded that homocysteine is a cardiometabolic biomarker with low sensitivity^24^. Currently, there are few articles in the literature showing this biomarker’s relevance as a tool for the prognosis of cardiometabolic risk in obesity.

Regarding the lipidic profile, significant statistic values were observed, being much higher in the obese groups compared to control groups, with an increase in total cholesterol, LDL-c, triglycerides, and a decrease in HDL. This fact is of great importance to the metabolic bias, for children and adolescents with these lipidic profiles may already exhibit fatty streaks in a coronary and carotid level, associated with the increase in CRP-us levels^25^. Suhet et al.^19^ observed the connection between CRP-us and low HDL. In a research conducted with 124 children, Muramoto et al.^26^ reported that for every increase of 1 mg/L in CRP serum concentration, a reduction of 0.072 mg/dL in HDL occurs, regardless of the individual nutritional state. When the LDL-c increases, it infiltrates the arterial endothelium, precociously producing fatty streaks around the first and second decade of life. The progression of dyslipidemia contributes to several subgroups of white cells infiltrating the vascular wall and secrete inflammatory cytokines, oxidative molecules, resulting in this inflammatory state. This mixed dyslipidemia, with posterior atheromatous plaques, was shown in Mexican children described by Gonzalez-Enriquez et al.^27^

According to Weiss et al.^28^, there is a positive correlation between the occurrence of obesity and dyslipidemia in children and adolescents. A prevalence of approximately 50% of dyslipidemia was found in children with high BMI, associated with a greater abdominal circumference, posterior accumulation of visceral fat, and non-alcoholic liver steatosis. Based on those findings, overweight is considered a sorting criterion of lipidic profile in obese children and adolescents, according to the Brazilian Pediatric Society (SBP) orientation^29^. Still, dyslipidemia during childhood may be associated with the early development of cardiometabolic risk with an increase in CRP-us, as observed in other populations of obese children and adolescents and the study sample^28,30^. In this sense, it can be concluded that the ultrasensitive C-reactive protein shows high sensitivity as a cardiometabolic risk biomarker in an obese pediatric population.

## Data Availability

The entire data set that supports the results of this study was published in the article itself.

## REFERENCES

1 Gupta N, Goel K, Shah P, Misra A. Childhood obesity in developing countries: Epidemiology, determinants, and prevention. Endocr Rev. 2012;33:48–70.

2 Fernandez RJ, Klimentidis YC, Keita AD, Casazza K. Genetic influences in childhood obesity: recent progress and recommendations for experimental designs. International Journal of Obesity. 2012;36:479–84.

3 Ezzati M. Worldwide trends in body-mass index, underweight, overweight, and obesity from 1975 to 2016: a pooled analysis of 2416 population-based measurement studies in 128·9 million children, adolescents, and adults. Lancet. 2017;390:2627–42.

4 Wabitsch M, Moss A, Kromeyer-Hauschild K. Unexpected plateauing of childhood obesity rates in developed countries. BMC Medicine 2014;12:1–5.

5 Rokholm B, Baker JL, Sørensen TIA. The levelling off of the obesity epidemic since the year 1999–a review of evidence and perspectives. Obes Rev 2010;1:835–46.

6 IBGE. Pesquisa de Orçamentos Familiares – POF 2008–2009: http://www.ibge.gov.br/home/estatistica/populacao/condicaodevida/pof/2008_2009_encaa/comentario.pdf. Acessado em 16/11/2010.

7 Melo ME. Os Números da Obesidade no Brasil: VIGITEL 2009 e POF 2008–2009.

8 Wang Z, Nakayama T. Inflammation, a link between obesity and cardiovascular disease. Mediators of Inflammation. 2010;173–86.

9 Junior AJS. Adipocinas: a relação endócrina entre obesidade e diabetes tipo II. Revista Brasileira de Obesidade, Nutrição e Emagrecimento. 2017;11:135–144.

10 Teixeira BC, Lopes AL, Macedo RCO, Correa CS, Ramis TR, Ribeiro JL, et al. Marcadores inflamatórios, função endotelial e riscos cardiovasculares. J Vasc Bras. 2014; 13:108–115.

11 Aday AW and Ridker PM (2019) Targeting Residual Inflammatory Risk: A Shifting Paradigm for Atherosclerotic Disease. Front. Cardiovasc. Med. 6:16. Doi: 10.3389/fcvm.2019.00016

12 Marshall JC, Reinhart K. Biomarkers of sepsis. Crit Care Med. 2009;37: 2290–9.

13 Han TS, Sattar N, Williams K, González, VC, Lean ME, Haffner SM. Prospective study of C-reactive protein in relation to the development of diabetes and metabolic syndrome in the Mexico City Diabetes Study. Diabetes Care. 2002;25:2016–21.

14 Guran O, Akalin F, Ayabakan C, Dereli FY, Haklar G. High-sensitivity C-reactive protein in children at risk for coronary artery disease. Acta Paediatrica. 2007; 96:1214–9.

15 Retnakaran R, Hanley AJG, Connelly PW, Harris SB, Zinman B. Elevated C-reactive protein in Native Canadian children: an ominous early complication of childhood obesity. Diabetes, Obesity and Metabolism. 2006;8:483–91.

16 Silva, LR, Stefanello JMF, Pizzi J, Timossi LS, Leite N. Aterosclerose subclínica e marcadores inflamatórios em crianças e adolescentes obesos e não obesos. Rev Bras Epidemiol 2012; 15(4): 804–16.

17 Roberts WL, Sedrick R, Moulton L, Spencer A, Rifoi N. Evaluation of four automated High sensitivity C-reactive protein methods: Implications for Clinical and Epidemiological Applications. Clinical Chemistry. 2000;46:461–6.

18 Kitsios K, Papadopoulou M, Kosta K, Kadoglou N, Papaggiani M, Tsiroukidou K. Highsensitivy c-reative protein levels and metabolic disorders in obese and overweight children and adolescentes. J Clin Res Pediatr En docrinol 2013;5(1):44–49. DOI: 10.4274/Jcrpe.789

19 Suhet LA, Hermsdorff HHM, Rocha NP, Silva MA, Filgueiras MS, Milagres LC, Peluzio, MCG, et al. Increased C-Reactive Protein in Brazilian Children: Association with Cardiometabolic Risk and Metabolic Syndrome Components (PASE Study). Cardiology Research and Practice. 2019;1–10.

20 Khera A, McGuire DK, Murphy SA, Stanek HG, Das SR, Vongpatanasin W, Wians FH Grundy SM, Lemos JA. Race and gender differences in C–reactive protein levels. J Am Coll Cardiol. 2005; 46:464–9.

21 Vasconcelos RS, Menezes CA. Clinical, dietary, and psychosocial effectiveness of interdisciplinary care in child and juvenile obesity. medRxiv doi: 10.1101/2020.06.09.20126953

22 Isasi CR, Deckelbaum RJ, Tracy RP, Starc TJ, Berglund L, Shea S. Physical fitness and C reactive protein level in children and young adults: The Columbia University BioMarkers study. Pediatrics. 2003;111:332–8.

23 Anderson JL, Muhlesteim JB, Horne BC, Carlquist JF, Madsen TE et al. Plasma homocysteine predicts mortality independently of traditional risck factors and C-reactive protein in patients with angiographically defined coronary artery disease. Circulation. 2000;102:1227–32.

24 Frauca JR, Gil EMG, Lozaro GB, Etayo PM, Martínez PV, López JPR, Lozano OB, Moreno LA. Abdominal fat and metabolic risk in obese children and adolescents. J Physiol Biochem. 2009; 65: 415–20.

25 Ridker, P.M. High-sensitivity C-reactive protein and cardiovascular risk: rationale for screening and primary prevention. Am J Cardiol. 2003; 92:17–22.

26 Muramoto G, Figueiredo Delgado A, Souza EC, A. Gilio E, Carvalho WB, Maranhão RC, Lipid profiles of children and adolescents with inflammatory response in a paediatric emergency department, Annals of Medicine. 2016;48:323–329.

27 Enríquez GVG, Benítez MIR, Gallegos VG, Buen EP, Sanromán RT, Cortés CLG. Contribution of TNF –308A and CCL2–2518A to carotid intima-media thickness in obese mexican children and adolescents. Archives of Med Research. 2008;39:753–9.

28 Weiss R, Dziura J, Burgert TS, Tamborlane WV, Taksali SE, Yeckel CW, et al. Obesity and the metabolic syndrome in children and adolescents. N. Engl J Med. 2004;350:2362–74.

29 Giuliano ICB, Caramelli B. Dislipidemias na infância e na adolescência. Pediatria. 2008;29:275–85.

30 Onis, M. Preventing childhood overweight and obesity. J. Pediatr (Rio J). 2015;91:105–7.

